# Magnitude and pattern of scabies among patients attending dermatology clinic of ALERT hospital; Addis Ababa, Ethiopia

**DOI:** 10.1101/2023.04.02.23287955

**Authors:** Betelhem Getachew, Amel Beshir, Elias Mulat

**Author notes:** Address of investigator.

## Abstract

**Background:** Scabies is a global public health problem and the highest prevalence of scabies occurs in tropical area, especially among marginal age groups and resource-limited communities. And it is one of the most common diseases seen in dermatology clinics.

**Objective:** To assess magnitude and pattern of scabies among patients attending dermatology clinic of alert hospital during the study period

**Method:** Hospital based retrospective study was conducted on scabies patients attending dermatology & venerology clinic, in ALERT hospital located in Addis Ababa, capital city of Ethiopia. The study population was all patients attending dermatology and venereology department at ALERT center between April 09,2015 to May 09, 2020. Data was collected about the number and demographic pattern of patients diagnosed with scabies from health management information systems (HMIS) registration book at dermatology clinic and sample of medical records of patients was reviewed by structured checklist about the clinical and treatment pattern of the disease.

**Result:** High frequency of scabies in patients presenting to a dermatology clinic is shown by this study. 5455 cases of scabies were registered from April 9,2015 to May 9,2020. 37226 total cases were seen in dermatology clinic of ALERT hospital between December 10, 2017 and November 11,2019. out of theis 37226 cases 2911 were scabies cases; making 7.8% of the outpatient visits by scabies patients.Of 5455 scabies cases registered, the majority (47.9%) belonged to the 15-45 years age group, and 26.8 % belonged to the 0-4 year age group. Those between 5-14 years of age consisted of 16.3 % of the total. Of the total scabies cases from the record, 3138 (57.5 %) were male and 2314 (42.4 %) were female. 1706 (28.9%) cases Kolfe keraniyo sub city, Addis Ababa and the other cases came to this hospital from different sub cities of Addis Ababa and rest of the country. The highest number of cases (n=1464) was registered during year September 12,2017 to September 10,2018. The median duration of symptoms for 25.7 % of the patients, is one month and three months for 17.3% (n=33) of them. Upper extremities were the most commonly affected body parts as mentioned on medical records of seventy patients. While the lower extremity genitalia and finger webs were mentioned as affected body parts following upper extremities. 87% of the lesions the patients were mentioned to be papules and the rest of the lesions constitutes of vesicle, plaque and Macule. 8 % (n=390) patients have had signs of infection. One patients diagnosis was mentioned as crusted scabies. Skin scraping was done for none of the patients. At ALERT hospital Benzyl benzoate was prescribed for 61.5% (n=249) of patients. 32.1% (n= 130) were given sulfur and 6.2% (n=25) were given permethrin. On this study,0.2% (n=1) patient was given ivermecin tablets. Permethrin soap was prescribed for four patients (0.9%) in addition to other drugs. 8 % (n=34) of the patients were given oral antibiotics because they had visible signs of infection. 7.8 % (n=33) were given anti pruritic agent. From a total of 71 medical records on which patients status after follow up was mentioned 62% (n=44) states the patient status as improved up on follow up. 23.9% (n=17) have the same disease status and 1.4 %(n=1) has worsened and 12.7(n=9) have post scabies itch. 0.04 % (n=16) from the total of 424 medical records revised have their diagnosis changed upon follow up and 0.07 % (n=29) have been retreated.

**Conclusion:** High frequency of scabies in patients presenting to a dermatology clinic is shown by this study.5455 cases of scabies were registered from April 9,2015 to May 9,2020. 37226 total cases were seen in dermatology clinic of ALERT hospital between December 10, 2017 and November 11,2019. out of these 37226 cases 2911 were scabies cases; making 7.8% of the outpatient visits by scabies patients.The highest proportion of the cases were male (57.5%) belonged to the 15-45 years age group and were widely from Kolfe keraniyo sub city (28.9%), Addis Ababa .Peak number of cases (n=1464) was registered from September 12,2017 to September 10,2018. The median duration of symptoms of scabies seen was one month. Upper extremities were mentioned as the most affected parts on the medical records of patients. Papules were the commonest lesion type mentioned on these records. Skin scraping was done for none of the patients. Benzyl benzoate was commonest drug prescribed. 62% patients status had improved up on follow up and 12.7% had post scabies itch.

## 1. Background

### 1.1 Introduction

Scabies is one of the most common dermatological problems affecting about 300 million people each year worldwide. It is caused by an infestation of the skin by the human itch mite (Sarcoptesscabiei var. hominis) (1,2).

The scabies mite usually is spread by direct, prolonged, skin-to-skin contact with a person who has scabies(1, 2)Also the infestation can possibly be spread by exposure to bedding, clothing as well as furniture that has been used(4,5). The incubation period is up to two months (2-6 weeks) for first time infestation; and 1-4 days for re infestation (2). Asymptomatic person with mites can also spread scabies (5).

Infestation with the scabies mite results in an intensely itchy skin eruption, which is most intense at night (5). The scabies symptoms involve almost all parts of the body but mostly affected sites include interdigital spaces, wrist, elbow, armpit, penis, nipple, waist, buttock, abdomen, shoulder places, feet, and thigh. In children and infants, head, feet, neck palms and soles are involved (3). In a small number of cases, although not exclusively, in the setting of immunosuppression, for example in those with advanced HIV infection and elderly, hyperinfestation can occur leading to crusted scabies, where the host may be colonized with many millions of mites. This is in contrast to classical scabies in which the host will harbour on average 10–15 mites (5).

The diagnosis of scabies is made largely on clinical grounds with typical distribution and characteristic visible serpiginous burrows. Handheld dermatoscope, parasitological confirmation with microscope can be used for diagnosis (5, 32).

A deep skin scrap is prepared using disposable scalpel and after disinfection of skin with 70% alcohol, wet mount smear will be prepared using 10% potassium hydroxide (KOH) to clear the specimen. The mite infestation is identified by observation of each form of Sarcoptes such as adult mites with spherical shape, four pairs of legs, hind legs end in long bristles, nymph or oval clear eggs with 0.1 - 0.15 mm long (35).

Scabies has a number of important sequelae. The severe, persistent pruritus which can be highly debilitating and stigmatizingand the itching being most intense at night is associated with sleep disturbance and a reduced ability to concentrate(5).The resultant scratching of the skin is an important cause of impetigo; most often due to Streptococcus pyogenes (group A streptococcus, GAS) and Staphylococcus aureus. Impetigo due to S. pyogenes acts as a precursor to invasive GAS infections, toxin-mediated diseases including scarlet fever and streptococcal toxic shock syndrome, and the autoimmune complications of rheumatic fever and glomerulonephritis (5, 32).

The highest burden of scabies is recorded in hot tropical and pacific countries. (3).Scabies is a major health problem in Africa specifically in the sub-Saharan region including Ethiopia(3).

In Ethiopia, scabies is common especially during natural or manmade disasters. For example, according to public health emergency measures surveillance report scabies is becoming beyond sporadic clinical cases but is turn to be a public health concern and affecting wider geographic areas and population groups especially in drought affected nutrition hotspot woredas(19). Scabies outbreaks in drought affected areas were seen in 2015 where there was shortage of safe water for drinking and poor personal hygiene as a result of direct impact of the drought caused by El Nino (19).

However, there is lack of studies regarding outbreak investigation and risk factors in the current study areas (19).In addition besides studies done on patterns of diseases in dermatologic clinic show scabies is one of most common cases seen(7,30) magnitude, clinical features and associated factors of scabies among patients attending health institutions is not well documented.

### 1.2 Statement of the problem

Scabies is a global public health problem and the highest prevalence of scabies occurs in tropical area, especially among marginal age groups and resource-limited communities.

The health consequences related to complications of scabies is one of the major problems especially in developing countries. In many countries, the main disabilities associated with scabies are caused by itching which could be enervating and stigmatizing and predisposes to secondary infection whichbrings a series of additional complications (27). Multiple factors such as poverty, poor hygiene, low socioeconomic conditions, overcrowded sleeping space, sharing of clothes have frequently reported as risk factors for scabies throughout the world(4).

As shown on reports done with the analysis of Records on the patterns of diseases in dermatologic clinics in Ethiopia scabies is one of the most common diseases (7,30).

Despite the disease burden, the magnitude, clinical features, factors associated with scabies in patients in Ethiopia visiting hospitals is not well documented (7,30). Therefore,studying magnitude and pattern of scabies helps to mitigate the disease burden, transmission; in addition to better the management of scabies in order to lessen its complications.

### 1.3 Significance of the study

Magnitude and distribution of cases by demographic factors and year and month of first visit is described in this study. This is done in order to help to point towards the source of cases to facilitate community interventions. In addition clinical, management patterns and treatment outcome are described.

Overall this study can provide important information for clinicians and researchers as well as policy makers.

## 2. Literature review

Scabies is a global public health problem and the highest prevalence of scabies occurs in tropical area, especially among marginal age groups and resource-limited communities (3). Scabies has global prevalence of 1.5% of the world population in 2010.(9) Scabies has been added to WHO ‘s list of Neglected Tropical Diseases (NTDs), in recognition of the very large burden of disease. However, unlike many other NTDs, scabies can also occur in temperate regions where it similarly has a predilection for vulnerable communities in which overcrowding and poverty coexist (12,4).

It is one of the commonest dermatological conditions which accounting for a considerable proportion of skin disease in developing countries (14,15). The complication is also associated with disabilities. According to the Global Burden of Disease (GBD) report, scabies directly accounts for 0.21% of global disability-adjusted life years (14). Scabies was seen in all age groups but showed a peak in children younger than 1 (19.9%), The greatest scabies burden, is highest in children aged 1–4 years and gradually decreasing from age 5 to 24 years (15).

Scabies affects almost all African countries. In sub-Saharan Africa the prevalence of scabies was ranged from 1-2 %(14).Drought and extreme water shortage is one of the driving forces of scabies outbreaks (16). 86% of the children in Sierra Leone displacement camps are affected by scabies and is high in countries with unstable conditions and civil war (3).

As a sub-Saharan Africa country; in Ethiopia Scabies is a major health problem (3). Since October 2015, scabies has been included as one of the reportable diseases in the drought affected areas. (17).In drought affected areas, the prevalence of scabies ranging from 2% to 67% (18). Moreover, in a recent systematic review, the prevalence of scabies ranged from 0.2% to 71.4% (2, 4). Likewise, on a study conducted in Northern Ethiopia among religious students; “Yekolotemeri” show that the overall prevalence rate was found to be 22.5 %(20).

Different risk factors for scabies have been described by different studies. Scabies affects people of all age groups; however, children in developing countries, elders, and immune compromised people are more vulnerable to scabies. (10, 11).

The prevalence of scabies was higher in rural areas than urban areas, 62.3% and 37.7%, respectively and it is more prevalent in cold and warmth seasons in males as well as females (21).

Also study in Cameroon the on prevalence of scabies among schools children scabies is linked with poor personal hygiene (22). The study done in Nigeria revealed that scabies is associated with poverty, overcrowding, and water scarcity (4,23). Scabies is more common in school children with a large family (≥7 members) (10.6%) as opposed to a family of 5-6 members (4.9%) and 3-4 members (3.8%) (24).

In the study done in outpatient clinic of dermatology in Egypt, Scabies occurred more frequent in those with family size of six (44.4%) members, having number of rooms less than four (96.3%), and with crowding index less than 2 (55.6%). The most patients with scabies were of low socioeconomic status (92.6%)(33).

On study done to assess burden of scabies in children of outpatient of Dhakashishur Hospital, in the capital of Bangladesh, 25.2% percent of the cases were found to have scabies clinically (6). In the outpatient clinic of dermatology in Egypt,out of 370 patients total of 27 cases were diagnosed as having scabies, giving a frequency rate of 7%(33).

A study done on Skin problems in children under five years old at a Gambo General Hospital, rural hospital in Southern Ethiopia; shows that the most common skin disease detected was scabies 13.6%. (30). According to another study done in Mekelle, Tigray, Ethiopia, from 2005 to 2009 with the analysis of Records of children and adolescents from the Italian Dermatological Centre, out of 17,967 medical records of outpatients and inpatients analyzed with patients aged 0 to 18 years, 2,777 had scabies.(25). From the ten leading causes of outpatient attendance on analysis of of medical records in Tigray, northern Ethiopia, scabies stands the fourth cause of the visits accounting for 7.3 % of the cases(7).

On a report on pattern of skin diseases at the University Teaching Hospital, Addis Ababa, from June 1995 to July 1997, only fourteen cases of scabies were seen out of 1505 patients. The low incidence of scabies in the referral clinic despite high incidence of scabies in the population was ascribed to the elimination of cases by internists and residents, and the early diagnosis and prompt treatment (8).

On the study done in Dhakashishur Hospital, Bangladesh; the patients were predominately male (male to female ratio of roughly 3;2(6). On the analysis of medical records in Tigray, northern Ethiopia, in the outpatient department (OPD) of the IDC, International Institute of Medical center, male to female ratio of scabies patients is 1.6.(7) According to a study done to assess factors associated with an outbreak of scabies at primary schools in southern Ethiopia 69.6% of the cases were male and 30.4% were female(3).

On the study done in Dhakashishur Hospital, Bangladesh; Toddlers were predominantly affected with the disease (35.1%) followed by infants (23.9%), preschool children (20.8%), olderchildren (12%) and adolescents (8.1%). Neonate were rarely affected with the disease (<0.1%) (6). In the outpatient clinic of dermatology in Egypt; the most common age in patients affected with scabies ranged from 30 to 49 years (33.3%), and 3.7% of the patients were aged more than 60 years(33).On the Mekelle, Italian Dermatological Centre study, scabies is the third most common dermatologic disease in children below six years of age.As per this study .Scabies was particularly common in younger children, probably because of closer contacts with parents or with other children. (25).

In the hospital based study of the outpatient clinic of dermatology in Egypt, itching increased at night in 96.3% of patients. And the most common presented clinical presentation was by scratch markings (70.4%). The most common site was hands and trunk (100% each), followed by axilla (92.6%), genitalia (81.48%), buttocks (70.37%), feet and legs (66.7% each), and breasts (7.4%) (33).

In a study done in Fiji, presence of both diseases in the scalp and face followed a clinical manifestation typical of infants and was rarely seen in older age groups. A possible explanation is the proximity of this body part to the mother ‘s breast or axillary areas while lactating (34).

A study conducted on outbreak of scabies at primary schools in southern Ethiopia, Damboya district has shown most common clinical symptom identified was itching skin rash which accounted for 99.17%. Nearly three-fourths, 73.41% of cases had tiny red borrows or blisters on their skin. Some of the cases had discharge from scabies skin lesion. The discharge from skin lesion is pus, 22.78% or blood, 1.26%. Crusted skin lesions were observed in 2.95% of the cases (3). As per a study done east Badewacho District, Southern Ethiopia 92% presented with itching sensation at night; almost all (92.7%) had rash on their finger and least (40%) on the anterior axillary and penis(9).

From the cases of scabies seen in outpatient department of Gambo General Hospital 44.3% of scabies were complicated with impetigo and/or eczema. (30). While none of the cases have sign of secondary infection in a study done on the out break of scabies east Badewacho District, Southern Ethiopia (9). On the study done in Dhakashishur Hospital, Bangladesh; about 60% percent of the cases were secondarily infected and 20% cases has eczematiztion change(6).People with scabies were 2.8 times more likely to have impetigo(34).scabies was the commonest cause of admission International Institute of Medical center in Mekelle, northern Ethiopia(7).

## 3. Objectives

### 3.1 General objective

- To assess magnitude and pattern of scabies among patients attending dermatology clinic of alert hospital during the study period

### 3.2 Specific objectives

- To determine the magnitude of scabies cases, among patients attending dermatology clinics of ALERT hospital during the study period.
- To determine type of distribution of scabies cases by demographic factors among patients attending dermatology clinics of ALERT hospital during the study period.
- To determine clinical, management patterns of scabies cases among patients attending dermatology clinics of ALERT hospital during the study period.

## 4 Methods and Materials

### 4.1 Study area and period

This study was conducted in All Africa Leprosy, Tuberculosis and Rehabilitation Training Centre (ALERT) located in Addis Ababa, capital city of Ethiopia, from May 2020-October 2020. ALERT is a medical facility focused on rehabilitation of leprosy patients, training programs on leprosy for personnel from around the world and leprosy control. The hospital currently provides a wide range of services in various departments including Dermatology, orthopedics and plastic surgery. It is one of the institutions where Addis Ababa university school of medicine dermatovenerology residency program is based. Currently it is also referral dermatology institute for dermatology cases from around Addis Ababa and all over Ethiopia. Even if most cases of scabies are first seen at the level of health centers, increasing number of cases are referred to this center for better diagnosis and management of cases.

### 4.2 Study design

Hospital based retrospective descriptive study design was used.

### 4.3 Study population

All patients attending dermatology and venerology department at ALERT center between April 9,2015 and May 9,2020.

### 4.4 Inclusion criteria

All cases of scabies, diagnosed by dermatology and venerology specialists on their first visits, documented in the medical records during past 5 years in ALERT hospital will be taken as eligible.

### 4.5 Exclusion criteria

Cases diagnosed with scabies on their follow up visits for other dermatologic.

### 3.6 Data collection and analysis

Data collection was started by collecting medical record numbers of patients diagnosed with scabies from health management information systems (HMIS) registration book at dermatology clinics. Overall 5455 medical record numbers, sex, age and address of patients with a diagnosis of scabies seen in the five year period (April 9,2015 to May 9,2020) was collected from the registration books.

Because of higher number of patients, all medical records cannot be reviewed for this study. Systematic random sampling was used.

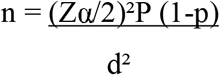

Where:

The sample size will be calculated as follows:

n= the required sample size, Zα/2 = Critical value of z= 1.96

P = period Prevalence of scabies among patients seen at dermatology OPD=25.2%

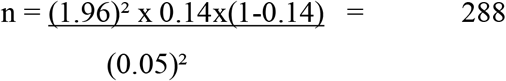

Additional 136 records were included because of incompleteness of the information on the records.

Patient ‘s medical records were reviewed and data was collected using structured questionnaire and analyzed using SPSS version 20.

### 4.7 Data quality assurance

Data extraction from health management information systems (HMIS) registration book and medical records of the patients was performed by the principal investigator. The data was checked manually to assess whether all available information are properly collected and recorded. Before processing, the collected data was checked for completeness.

### 4.8 Ethical considerations

Letter of support obtained from the dermatovenereology department was submitted to ALERT Hospital for permission to collect data from patient medical records. Anonymity and confidentiality of the patient information was kept private. The name of the patients was not included in the data collection format and their medical record number was not included in the analysis.

## 5. RESULTS

### 5.1 Frequency of scabies cases

High frequency of scabies in patients presenting to a dermatology clinic is shown by this study. 5455 cases of scabies were registered from April 9,2015 to May 9,2020. 37226 total cases were seen in dermatology clinic of ALERT hospital between December 10, 2017 and November 11,2019. out of theis 37226 cases 2911 were scabies cases; making 7.8% of the outpatient visits by scabies patients.

### 5.2 Distribution of cases by socio demographic status

Of 5455 scabies cases registered, the majority (47.9%) belonged to the 15-45 years age group, and 26.8 % belonged to the 0-4 year age group. Those between 5-14 years of age consisted of 16.3 % of the total. Of the total scabies cases from the record, 3138 (57.5 %) were males and 2314 (42.4 %) were females. 1706 (28.9%) cases Kolfe keraniyo sub city, Addis Ababa and the other cases came to this hospital from different sub cities of Addis Ababa and rest of the country as specified on the table. (Table 1)

**Table 1.**
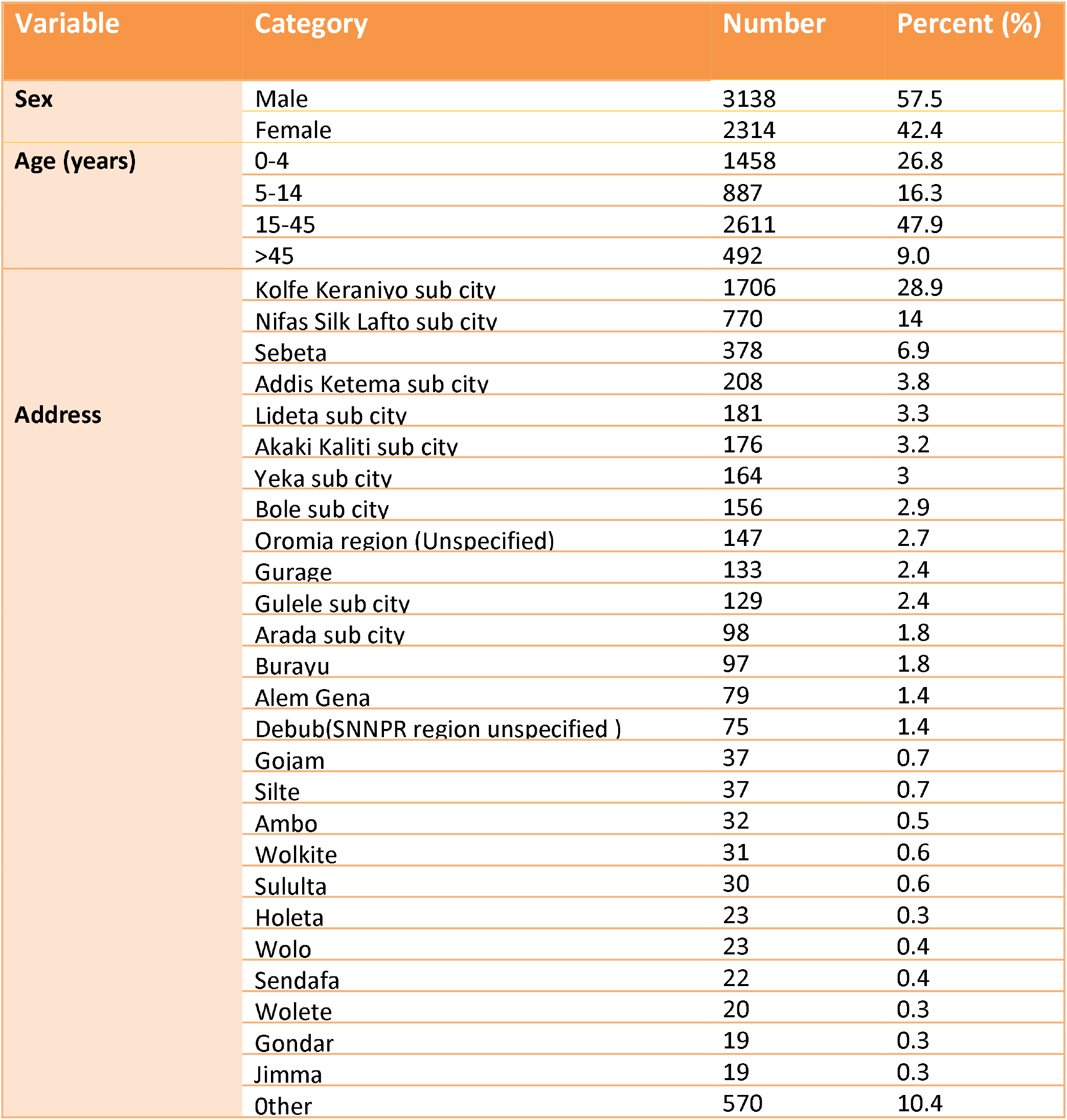
Distribution of cases by sociodemographic status (Age, Sex, Address)

**Figure 1.**
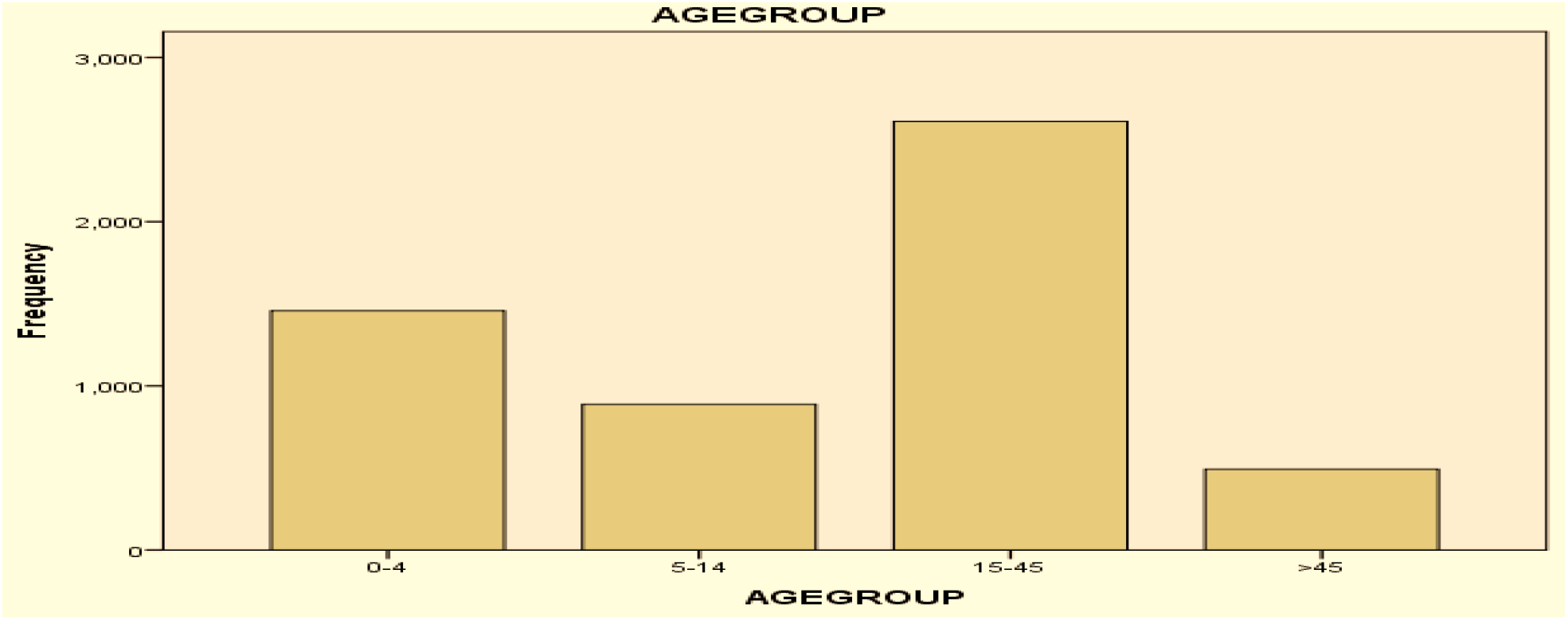
Distribution of cases by age.

**Figure 2.**
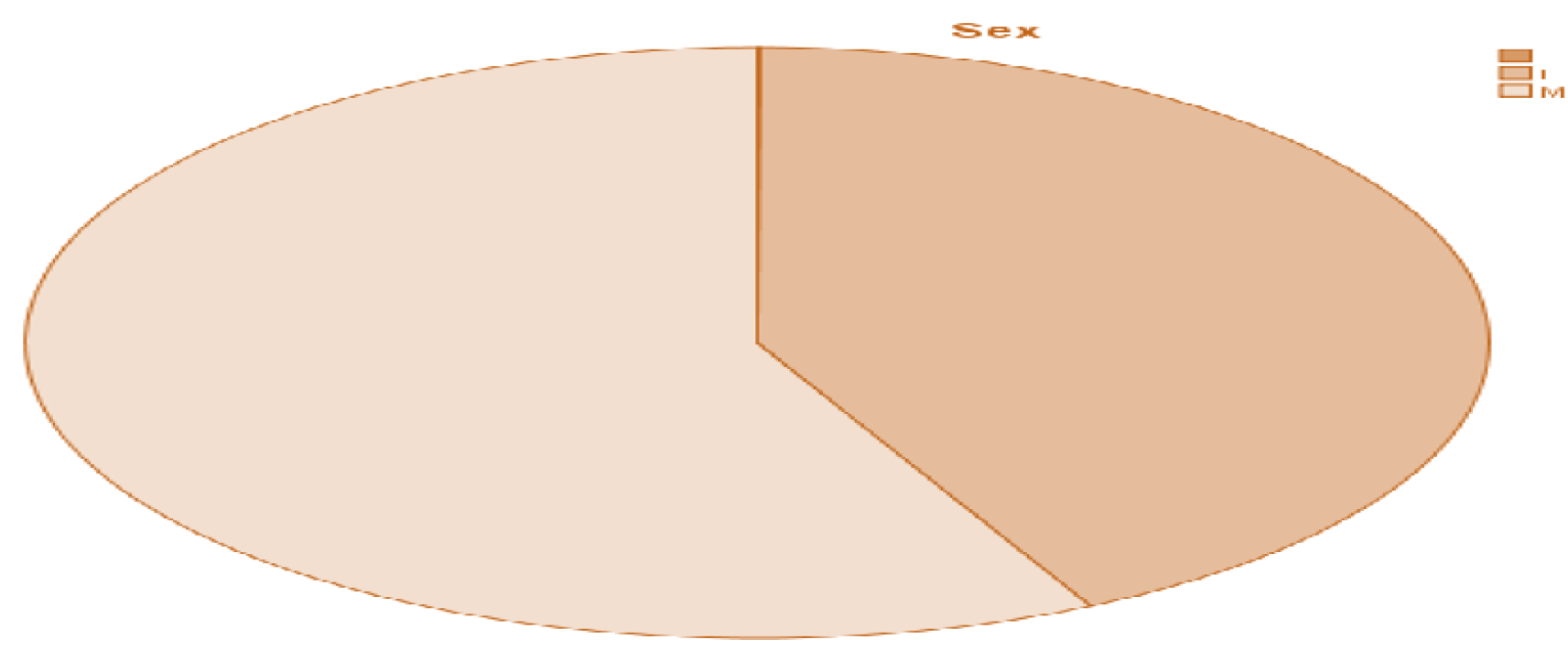
Distribution of cases by sex.

### 5.3 Distribution of Cases by Year and Month

The highest number of cases (n=1464) was registered from September 12,2017 to September 10,2018. (Table 2).

**Table 2.**
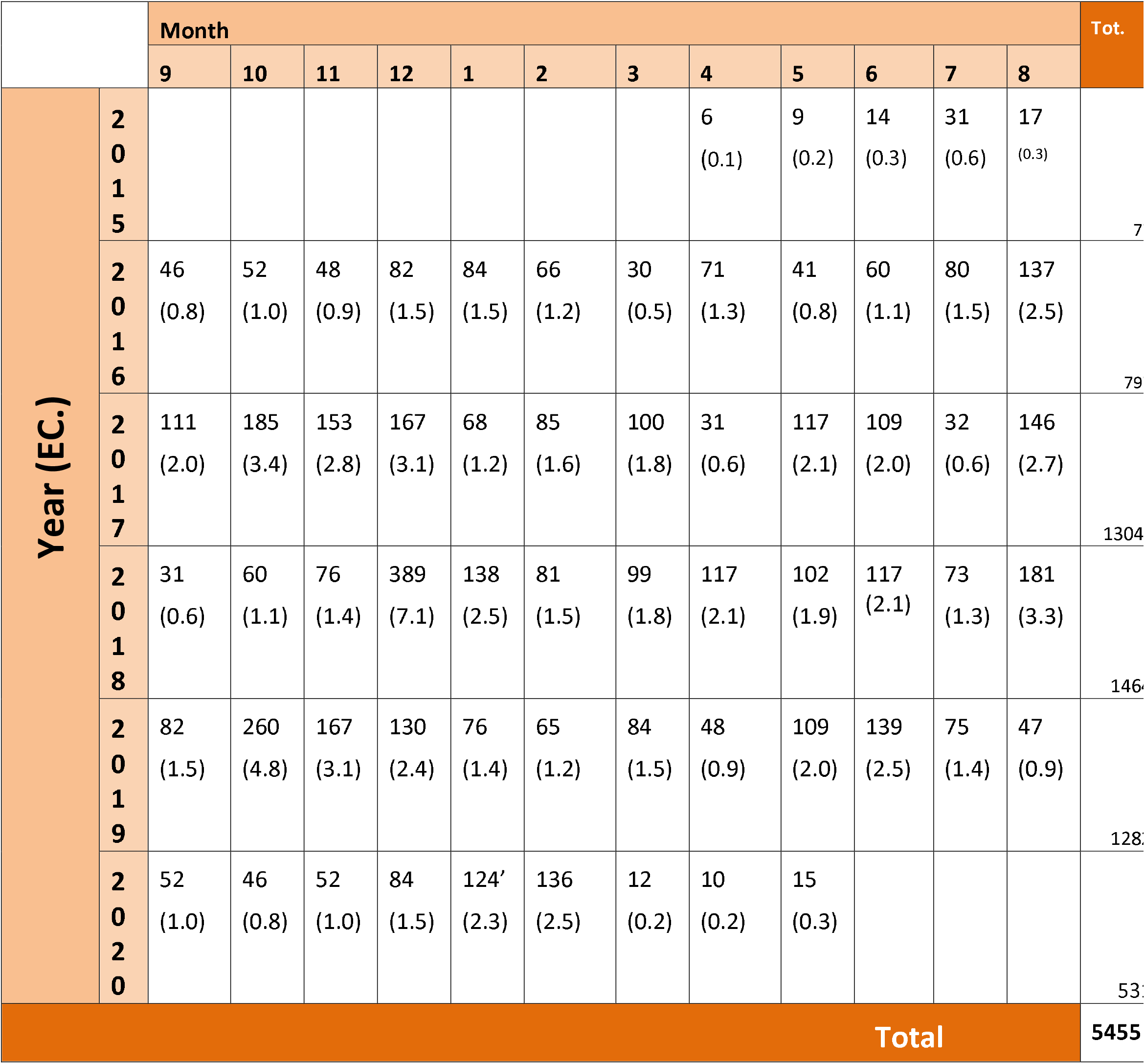
Distribution of number of cases (percent) by year and month of patient ‘s first visit

**Table 3.**
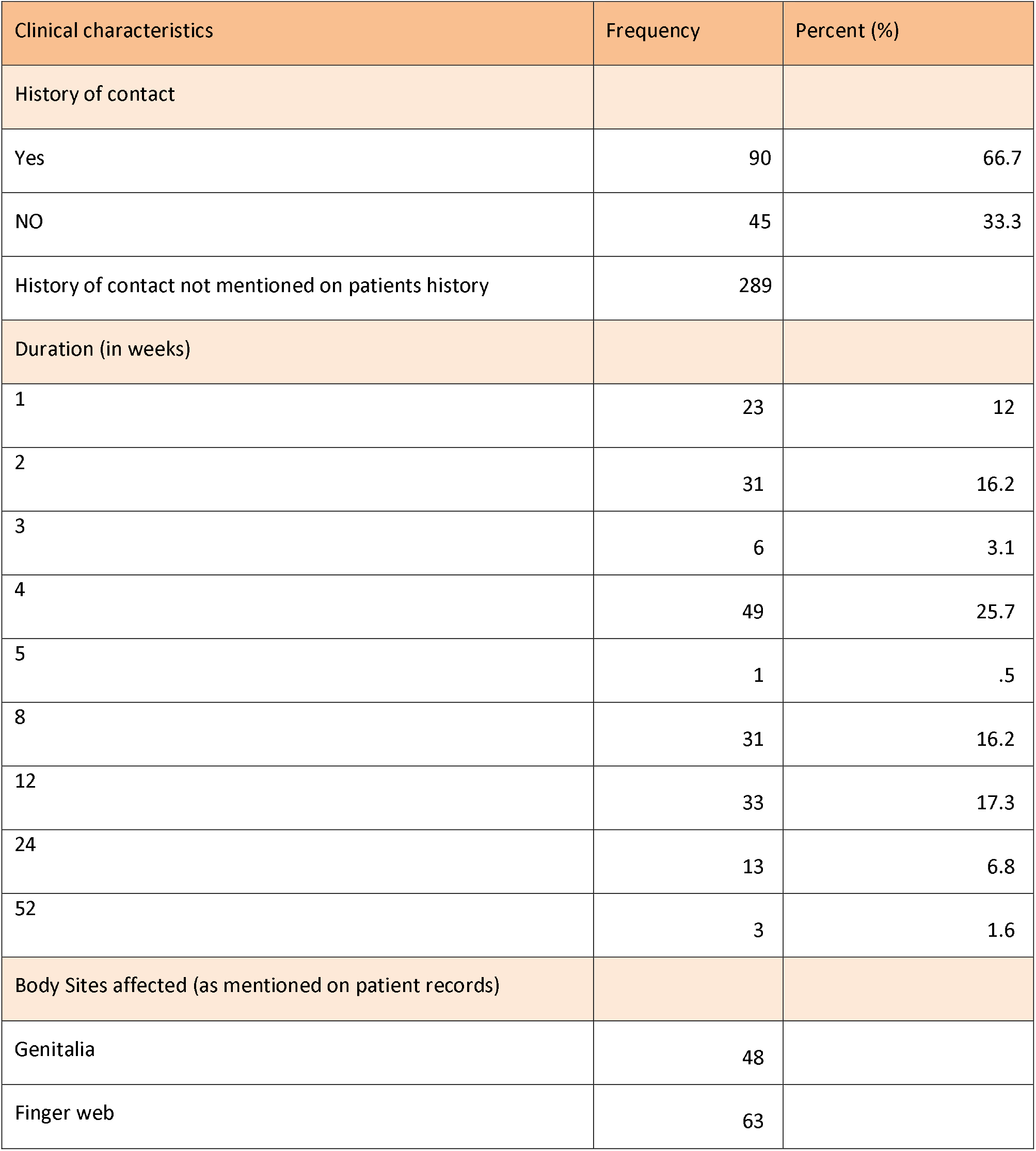
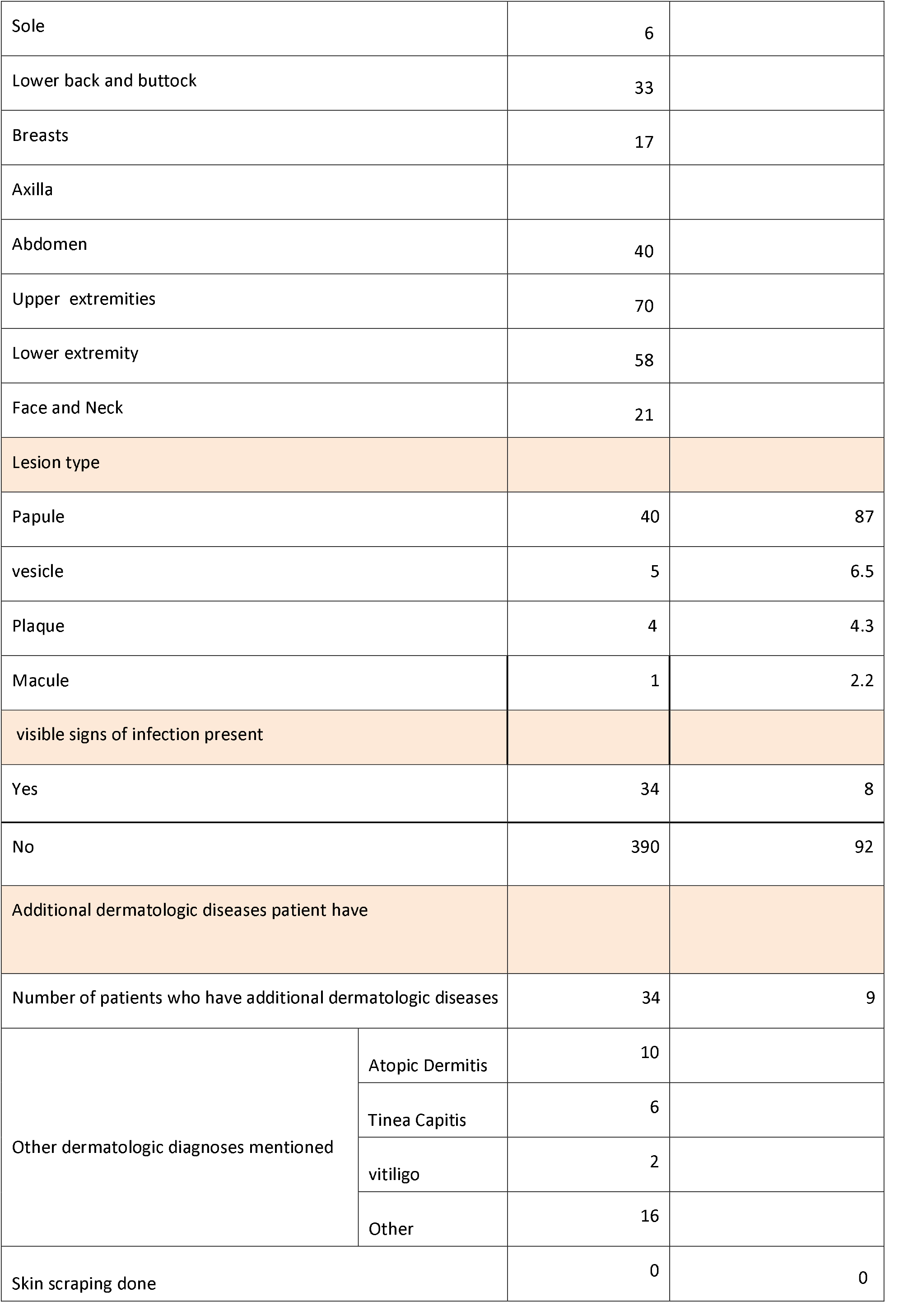
Distribution of cases by clinical characteristics

### 5.4 Distribution of cases by clinical characteristics

The median duration of symptoms, 25.7 % of the patients, is one month and 17.3% (n=33) of them for three months (12 weeks). The involvement of Upper extremities was mentioned on medical records of seventy patients. While the lower extremity genitalia and finger web were mentioned as affected body parts on fifty eight, forty eight and sixty three patients medical records consecutively.87% of the lesions the patients have were mentioned to be papules and the rest of the lesions constitutes of vesicle, plaque and Macule. 8 % (n=390) patients have had signs of infection.One patient ‘s diagnosis was mentioned as crusted scabies.

Skin scraping was done for none of the patients.

### 5.5 Treatment history at other health facilities

Treatment for the same complaint at other health facilities was mentioned on sixteen patient ‘s history for which three of the cases were given topical steroids and two cases were treated with Sulfur 5% and the rest of the cases were given unspecified medications. All cases had no improvement with the above mentioned treatments at the facilities.

### 5.6 Types and frequencies of antiscabious, antipruritics and antibiotics prescribed

Benzyl benzoate was prescribed for 61.5% (n=249) of patients. 32.1% (n= 130) were given sulfur and 6.2% (n=25) were given permethrin. On this study,0.2% (n=1) patient was given ivermecin. Permethrin soap was prescribed for four patients (0.9%) in addition to other drugs.(Table 4) 8 % (n=34) of the patients were given per oral antibiotics because they had visisble signs of infection. 7.8 % (n=33) were given anti pruritic agent.

**Table 4.**
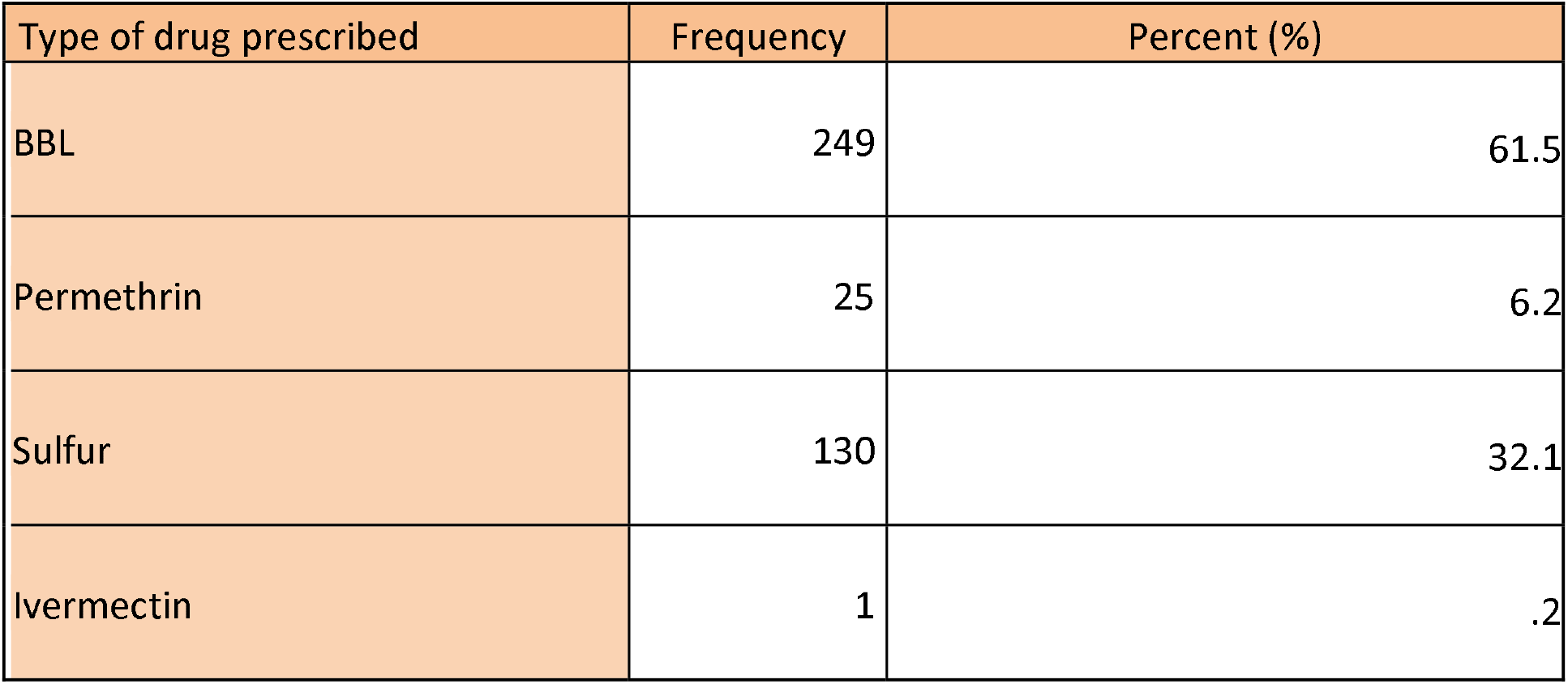
Types and frequencies of Antiscabious drugs prescribed

| Type of drug prescribed | Frequency | Percent (%) |
| --- | --- | --- |
| BBL | 249 | 61.5 |
| Permethrin | 25 | 6.2 |
| Sulfur | 130 | 32.1 |
| Ivermectin | 1 | .2 |

### 5.7 Treatment out come after administration of antiscabiuos drugs

From a total of 71 medical records on which patients status after follow up was mentioned 62% (n=44) states the patient status as improved up on follow up. 23.9% (n=17) have the same disease status and 1.4 %(n=1) has worsened and 12.7(n=9) have post scabies itch. 0.04 % (n=16) from the total of 424 medical records revised have their diagnosis changed upon follow up and 0.07 % (n=29) have been retreated. (Table 5 and 6)

**Table 5.1.** Treatment out come after administration of antiscabiuos drugs

| Status of the patient on follow up | Frequency | Percent (%) |
| --- | --- | --- |
| Improved | 44 | 62.0 |
| same | 17 | 23.9 |
| worsens | 1 | 1.4 |
| Post scabies itch | 9 | 12.7 |
| Total | 71 | 100.0 |

**Table 5.2.** Frequency and percentage of patients with diagnosis change and retreatment after treatment for scabies

| Status of the patient on follow up | Frequency | Percent (%) |
| --- | --- | --- |
| Diagnosis changed | 16 | 0.04 |
| Retreated | 29 | 0.07 |

**Figure 4.**
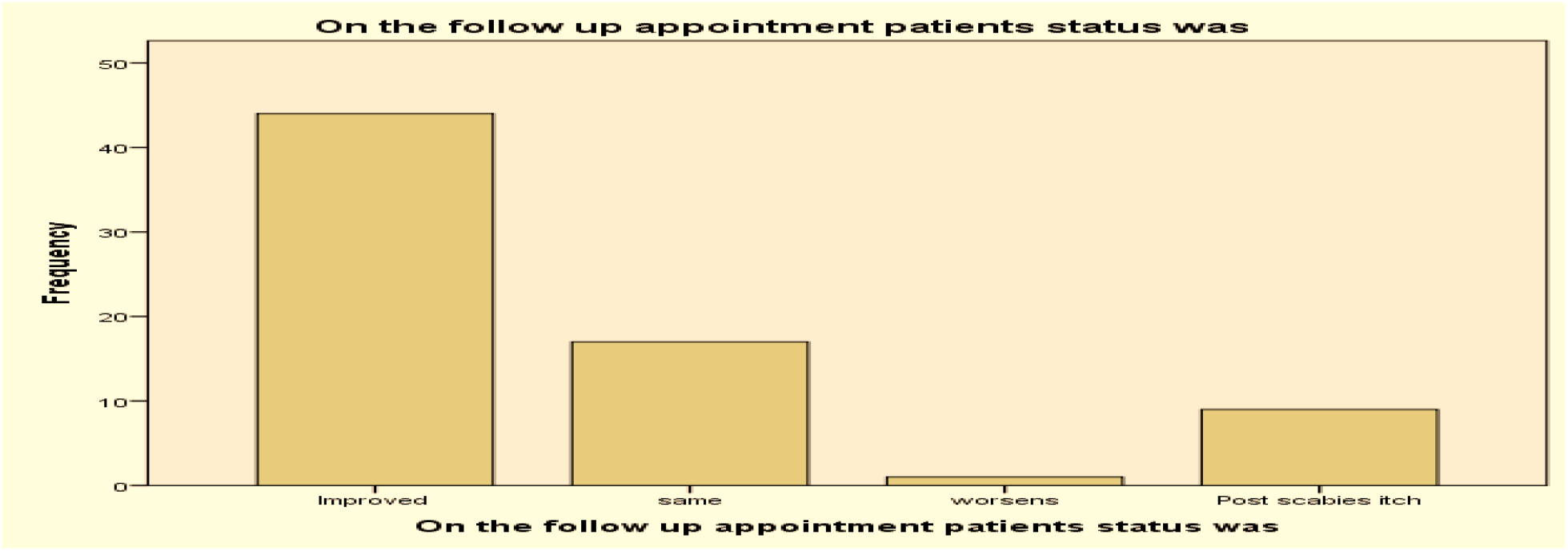
Treatment out come after administration of antiscabiuos drugs.

## 6. Discussion

Scabies is a common problem in Ethiopia with a few outbreak investigations. High frequency of scabies in patients presenting to a dermatology clinic is shown by this study. out of 37226 cases seen in dermatology clinic of ALERT hospital between December 10, 2017 and November 11,2019. 2911 were scabies cases; making 7.8% of the outpatient visits by scabies patients.

This is in accordance with the analysis of medical records in Tigray, northern Ethiopia, in the outpatient department (OPD) of the IDC, International Institute of Medical center, Scabies comprises 7.3 % of outpatient visits.(7)

A study done on Skin problems in children under five years old at a Gambo General Hospital, rural hospital in Southern Ethiopia; shows that the most common skin disease detected was scabies 13.6%. (30).In the outpatient clinic of dermatology in Egypt, out of 370 patients total of 27 cases were diagnosed as having scabies, giving a frequency rate of 7%(33).

But on a report on pattern of skin diseases at the University Teaching Hospital, Addis Ababa, from June 1995 to July 1997, only fourteen cases of scabies were seen out of 1505 patients. The low incidence of scabies in the referral clinic despite high incidence of scabies in the population was ascribed to the elimination of cases by internists and residents, and the early diagnosis and prompt treatment (8).

Out of 5448 scabies cases registered; the majority, 47.9%(n=2611), belonged to the 15-45 years age group, and 26.8%(n=1458) belonged to the 0-4 years age group. Those between 5-14 years of age consisted of 16.3 % of the total. The mean age of patients was 20 +/-18 years on this study. In study done in the outpatient clinic of dermatology in Egypt; the most common age affected ranged from 30 to 49 years (33.3%), and 3.7% of the patients were aged more than 60 years(33). In study done in Tikrit, a city found in IRAQ highest prevalence was in children [15.6%], followed by the age group 30 to 44 years [14.7%].(IRAQ)

On the Mekelle, Italian Dermatological Centre study, scabies is the third most common dermatologic disease in children below six years of age. (25). In community based study done in Kechabira district, Southern Ethiopia the most affected age group were those in the age group of 5–14 with an attack rate of 131/1000 population. Analogous fndings were recorded by investigations East Badewacho District, Southern Ethiopia. (19) The possible reasons for wide spread of scabies among young children could be the close contact among peers, overcrowding in schools and sharing of contaminated private materials particularly closes.(19)

The male to female ratio in this study is 1.4 which is close to the result seen on the analysis of medical records in Tigray, northern Ethiopia, in the outpatient department (OPD) of the IDC, International Institute of Medical center, which is 1.6 (7). On the study done in Dhakashishur Hospital, Bangladesh; the patients were predominately male (male to female ratio of roughly 3:2(6). The attack rate of scabies among both sexes was nearly the same in Scabies outbreak investigation in Kechabira district, Southern Ethiopia (36).

According to a study done to assess factors associated with an outbreak of scabies at primary schools in southern Ethiopia 69.6% of the cases were male and 30.4% were female (3).

The majority of patients, 28.9 % (n=1706) came from Kolfe keraniyo kifle ketema, Addis Abeba and 14% (n=768) came from Nifas Silk Lafto Kifle ketema. 378(6.9%) patients came from Sebeta, Oromia region constituting third highest number of patients.

The highest number of cases was registered during the year 2010 E.C, followed by 2009 and 2011. The number of patients shows significant decrements because of corona pandemic since seventh month of the Ethiopian calendar year 2012.

With reference to the sites of lesions, the involvement of Upper extremities was mentioned on medical records of seventy patients. While the lower extremity genitalia and finger web were mentioned as affected body y parts on fifty eight, forty eight and sixty three patients medical records consecutively.

This is in accordance with study done in Fiji where lesions were most often observed on the upper limbs (34). In the hospital based study of the outpatient clinic of dermatology in Egypt, most common site was hands and trunk (100% each), followed by axilla (92.6%), genitalia (81.48%), buttocks (70.37%), feet and legs (66.7% each), and breasts (7.4%) (33)

87% of the lesions the patients have were mentioned to be papules and the rest of the lesions constitutes of vesicle, plaque and Macule. This is similar with study of The epidemiology of scabies in Gaza Governorates, where patients presented with papules (53%), vesicular lesions (37%), nodules (5.9%), and erythema (2.9%)(38).

One patient ‘s diagnosis was mentioned as crusted scabies. But in a study done on factors associated with scabies outbreaks in primary schools in Ethiopia Crusted skin lesions were observed in 2.95% (7/237) of the cases(3).

8 % (n=34) patients have had signs of infection. But this number is relatively less than the cases of scabies seen in outpatient department of Gambo General Hospital 44.3% of scabies were complicated with impetigo and/or eczema. (30). While none of the cases have sign of secondary infection in a study done on the out break of scabies east Badewacho District, Southern Ethiopia (9). On the study done in Dhakashishur Hospital, Bangladesh; about 60% percent of the cases were secondarily infected (6). The relatively smaller number of secondary infection could be due to treatment at health centers before referral.Benzyl benzoate was prescribed for 61.5% (n=249) of patients. 32.1% (n= 130) were given sulfur and 6.2% (n=25) were given permethrin. On this study, 0.2% (n=1) patient was given ivermecin. Permethrin soap was prescribed for four patients (0.9%) in addition to other drugs. On study done regarding factors associated with scabies outbreaks in primary schools in Ethiopia among patients that had taken medical treatment, 16.03% (34/237) used permethrin 5% lotion and 0.84% (2/237) benzyl benzoate lotion(3).

From a total of 71 medical records on which patients status after follow up was mentioned 62% (n=44) states the patient status as improved up on follow up. 23.9% (n=17) have the same disease status and 1.4 %(n=1) has worsened.On a done Survey in a Dermatology Clinic in Iraq, topical application of 5% permethrin cream demonstrated a cure rate of 80.3% following a single application. The cure rate increased to 95.5% after a second application and complete cure was achieved for all cases after the third application. (37)

## 7 Conclusion and recommendation

### 7.1 Conclusion

High frequency of scabies in patients presenting to a dermatology clinic is shown by this study 7.8% of the outpatient visits of dermatology clinic of ALERT hospital December 10, 2017 and November 11,2019.

The highest proportion of the cases were male (57.5%) belonged to the 15-45 years age group and were widely from Kolfe keraniyo sub city (28.9%), Addis Ababa .Peak number of cases (n=1464) was registered from September 12,2017 to September 10,2018 .

The median duration of symptoms of scabies was one month .

Upper extremities were mentioned as the most involved parts on medical records. Papules were the commonest lesion type mentioned. Skin scraping was done for none of the patients. Benzyl benzoate was commonest drug prescribed; for 61.5% of patients. 62% patient ‘s status has improved up on follow up. 23.9% have the same disease status and 1.4 % has worsened and 12.7% have post scabies itch. Over all, some of the findings in this study are in line with previous reports from community and hospital based studies in Ethiopia and other countries.

### 7.2 Recommendation

- Oral ivermectin is an effective and safe alternative especially for mass treatment.
- Health professionals working in health centers should further strength their knowledge on the assessment and treatment of complications, and consultation on contact and fomite treatment.
- Health education prevention and control with emphasis on keeping personal hygiene should be worked up on.
- Additional community and hospital based studies has to be carried out in the future to better perceive the epidemiology impact of scabies.

## 8. Limitations of the study

- Diagnosis was carried out only on the clinical basis; not confirmed by burrow scraping and microscopic examination.
- Incomplete history and physical findings in patient medical records have affected the data quality.
- As a hospital-based study, this study may overestimate or underestimate the true incidence of the disease.

## Data Availability

All data produced in the present study are available upon reasonable request to the authors
All data produced in the present work are contained in the manuscript.

## ACKNOWLEDGEMENT

I would like to thank Addis Ababa University; school of medicine, Dermatology and venereology department for creating and supporting this great opportunity of research works.

## Abbreviations

A.A: Addis Ababa
AAU: Addis Ababa university
ALERT: All African Leprosy Rehabilitation and Training center
FMOH: Federal Ministry of Health
WHO: World Health Organization
E.C: Ethiopian Calendar
G.C: Gregorian Calendar
GAS: Group A streptococcus
HIV: Human immune deficiency virus
HMIS: Health Management information systems
KOH: potassium hydroxide

## 10. Annex

### Data collection format

01. Questioner serial no_________________________date______________

02. Medical record no.____________________________________________

03. Address- Region______Town ____________k/ketema_____________

04. Sex 1. Male 2.Female

05. Age_________________________

06. Symptoms: 1.Skin rash 2. Night itching 3. Other

07. Duration:________________________________________

08. Itching is intense during the: 1. Day 2. Night

09. Contact history 1.yes 2. No

10. Hx of Rx for same complaint at other health facilities :1. Yes 2. No

11. Mention the drug given at other health facility:_______________________

12. Is lesion visible? 1. Yes 2. No

13. Lesion:1. Vesicles 2. Papule 3. 4. Nodule 5.Pustules and crust 6. Other

14. Body parts affected ______________________________________-

15. Variant of scabies____________________________________

16. Presence of visible signs of infection: 1. Yes 2. No

17. Is the lesion crusted scabies? 1.Yes 2.No

18. Immunosuppression: 1. Yes 2.No

19. Other skin condition: 1.yes 2. No

20. Mention the skin condition__________________________________

21. Required admission: 1. Yes 2. No

22. Antibiotic prescribed: 1. Yes 2. No

23. Drug prescribed

24. Anti-pruritic prescribed 1.yes 2. No

25. Anti pruritic :___________________________________

26. Skin scraping done 1.yes 2.No

27. If yes : 1. Positive 2. Negative

28. Contact treatment: 1. Yes 2. No

29. Appointment was given : 1. Yes 2.No

30. Appointed after:___________________________________________

31. On follow up: 1. Improved 2. Same 3. Worsens 4. Post scabies itch

32. Diagnosis changed : 1.yes 2.No

33. Retreated: 1.yes 2.No

